# Catheter Laboratory Facilities in Indonesia: Number, Growth, and Distribution in The Largest Archipelago Nation

**DOI:** 10.1101/2023.04.14.23288607

**Authors:** Farizal Rizky Muharram, Andrianto Andrianto, Senitza Anisa Salsabilla, Chaq El Chaq Zamzam Multazam, Wigaviola Socha Harmadha, Iwan Dakota, Hananto Andriantoro, Doni Firman, Radityo Prakoso

## Abstract

**Background:** Indonesia, the world’s largest archipelago and fourth most populous nation, has limited transportation due to geographical obstacles. This affects the provision of acute time-dependent therapy such as Percutaneous Coronary Intervention (PCI). Indonesia’s ageing population, which will increase Acute Coronary Syndrome prevalence in the next decade, will worsen this problem. Therefore, the analysis and enhancement of cardiovascular care are crucial. The catheterization laboratory (cath lab) performs PCI procedures. This study maps the number and distribution of Indonesia’s cath lab facilities.

**Methods:** A direct survey was used to collect the cath lab location data. Population data came from the Ministry of Home Affairs. The growth of cath labs is shown and analyzed by region. The ratio and the Gini index are the primary comparison tools between regions and over time.

**Findings:** The number of cath labs in Indonesia significantly increased from 181 to 310 in the last five years, with 44 of the 119 new labs built in an area that did not have one. Java has the most cath labs (208, 67%). The cath lab ratio in the provinces of Indonesia ranges from 0·0 in West Papua and Maluku to 4·46 in Jakarta; the median is 1·09. (IQR 0·71–1·18). The distribution remains a problem, as shown by the high cath lab Gini index (0·48).

**Interpretation:** The number of cath labs in Indonesia has increased significantly recently. However, maldistribution remains a concern. In order to improve Indonesia’s cardiovascular emergency services, Future cath lab development must be planned better by considering the facility accessibility and density.

**Funding:** The study was conducted with the researcher’s funds

**Evidence Before Study:** Indonesia is the fourth-largest nation by population and the largest archipelagic country. Unfortunately, the number of cath lab facilities nationwide in Indonesia was never examined or mentioned in any previous scientific articles that we could find. According to earlier studies, cathlab accessibility is critical in reducing the time ACS patients take to receive care. It is become crucial to map cathlab locations and plan them in strategic locations.

**What this Study Adds:**

- This study provides data on the number of cathlabs, their primary geographic distribution, their cath lab-to-population ratio, and their evolution over the previous five years in Indonesia.
- Our research demonstrates that to ensure equity access, Indonesia, as the largest archipelagic nation, needs government policies that initiate the distribution of cathlabs and the strategic placing of cathlabs as critical factors.

## Background

Indonesia has entered a new population growth phase, and its demographic distribution will shift to an older population in the next ten years. It is estimated that the population will increase by 6–7% from 2020 to 2030, with the elderly population (>45 years old) increasing by up to 30%, a rate four times greater than that of the general population. This will lead to an increase in the incidence of cardiovascular disease, particularly acute coronary syndrome.

Acute Coronary Syndrome (ACS) is a life-threatening noncommunicable disease. Its effective treatment depends upon specific and general infrastructure. Specific infrastructure includes health facilities with specialized equipment (cath lab), and general infrastructure (roads, ambulances, referral systems) supports access to these facilities. Primary Percutaneous coronary intervention (PCI) is still the gold standard in treating ACS as evidenced by a reduction in mortality and morbidity in patients receiving this treatment. However, the efficacy of primary PCI depends upon symptoms-to-needle and door-to-needle time criteria. ^1–3^

As one of the world’s largest archipelago countries, Indonesia has the burden of a growing population, and a transportation and access problem. In addition, as an archipelago, the sea is a barrier between islands, separating their populations and making it harder for health facilities to cover their people. Access to health facilities needs improved transportation. In this paper, we will describe the growth, number, ratio, and distribution of cath labs in Indonesia with the expectation of it providing a basis for policy-making.^4–6^

## Methods

### Conceptual framework

This study is cross-sectional, with geospatial information systems used for the primary analysis. We gathered the data on the cath lab facilities, road networks, population, and demographic boundaries and analyzed them using Network Analysis. This study was divided into two phases: The first phase was data collection, and the second was data analysis. The process will be held in July 2022. In this research, we focused on the infrastructure aspect, such as the road network and cath lab facility. This paper does not discuss other factors that may affect the time needed to reach the nearest facility, such as the cardiac emergency system, human resources available to perform PCI, and the national referral system [Figure 1].

**Figure 1.**
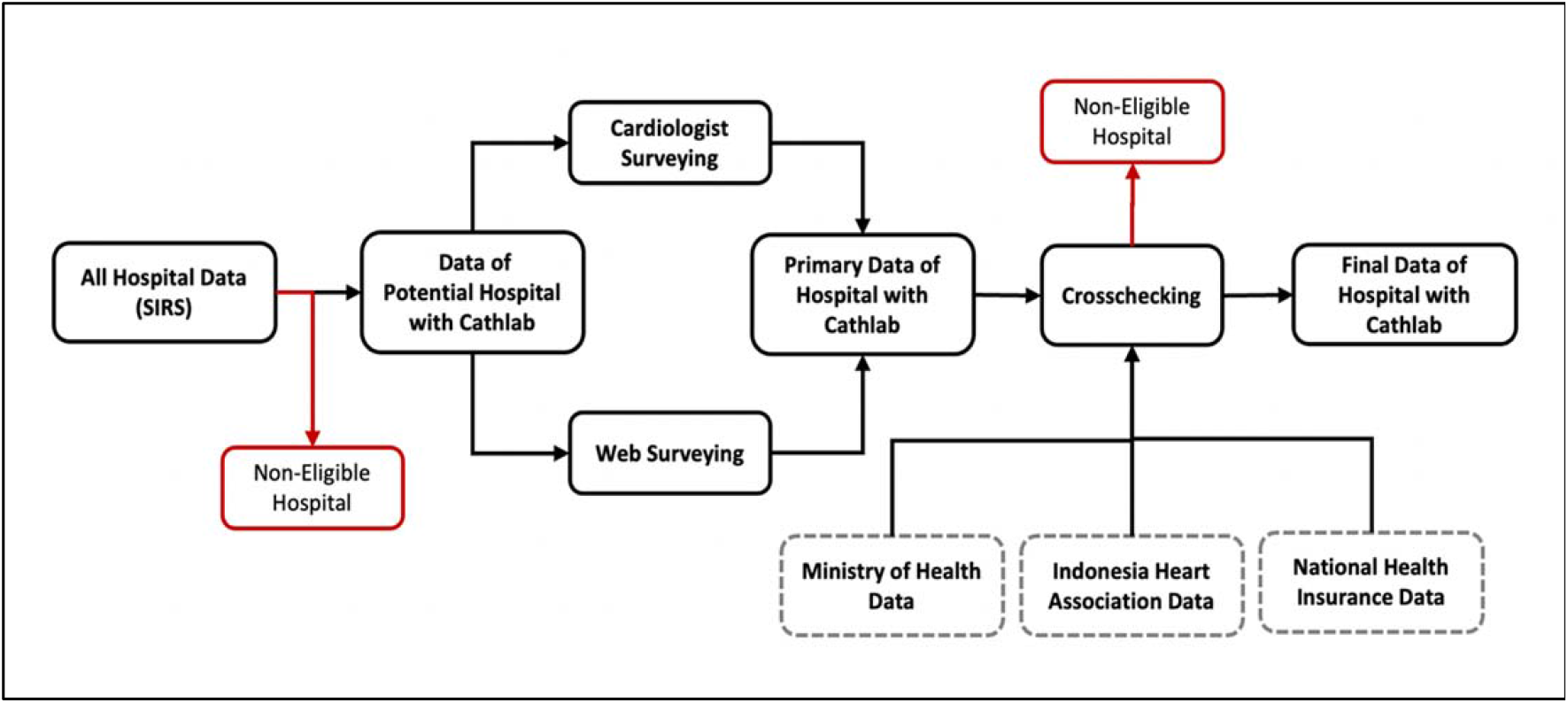
Cath lab data collecting process.

**Figure 2.**
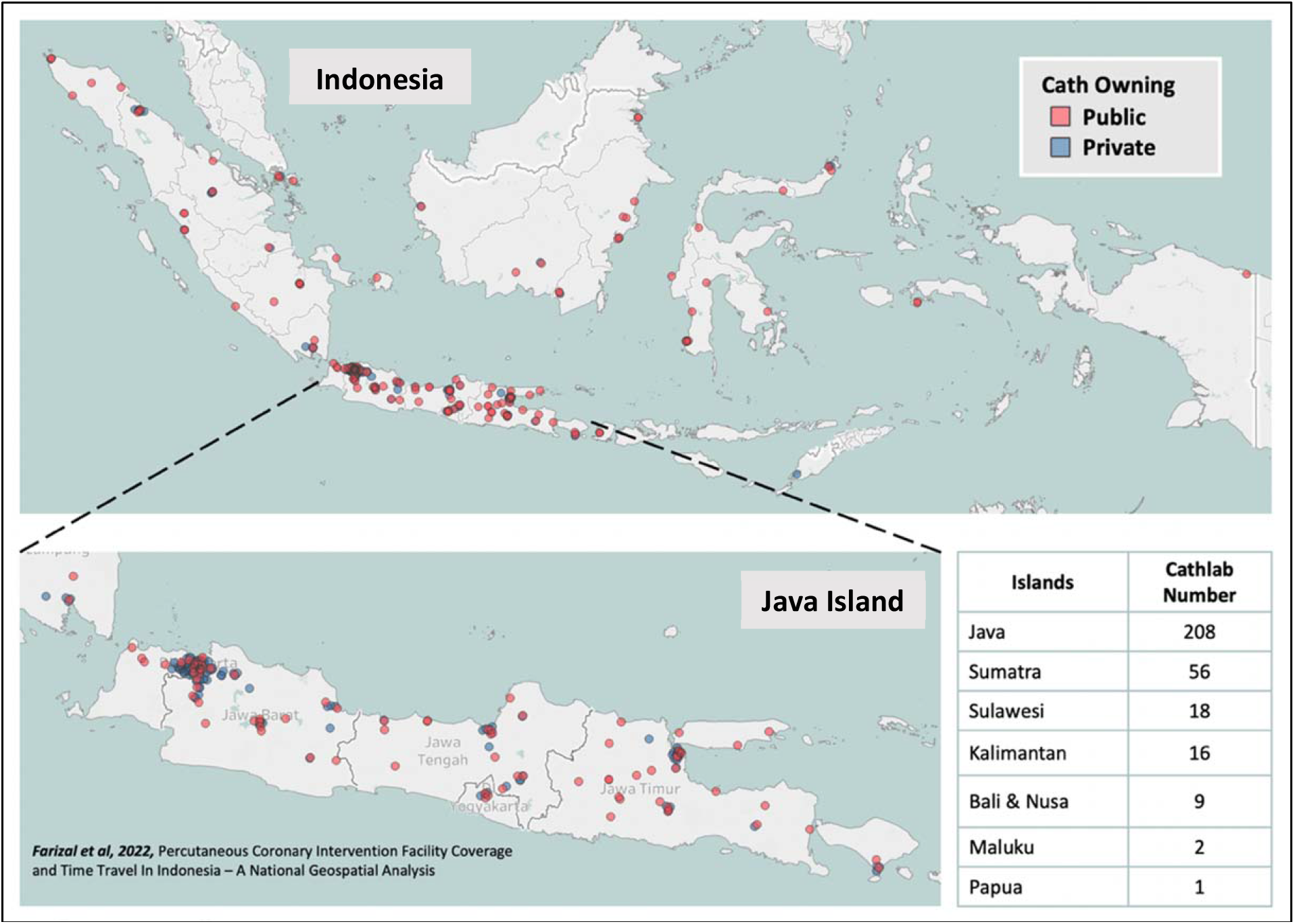
The map of Indonesia cath lab distribution, the red dot represents cath lab owned by public hospitals, and blue by private hospitals. The table in lower right shows the distribution between the major islands in Indonesia.

**Figure 3.**
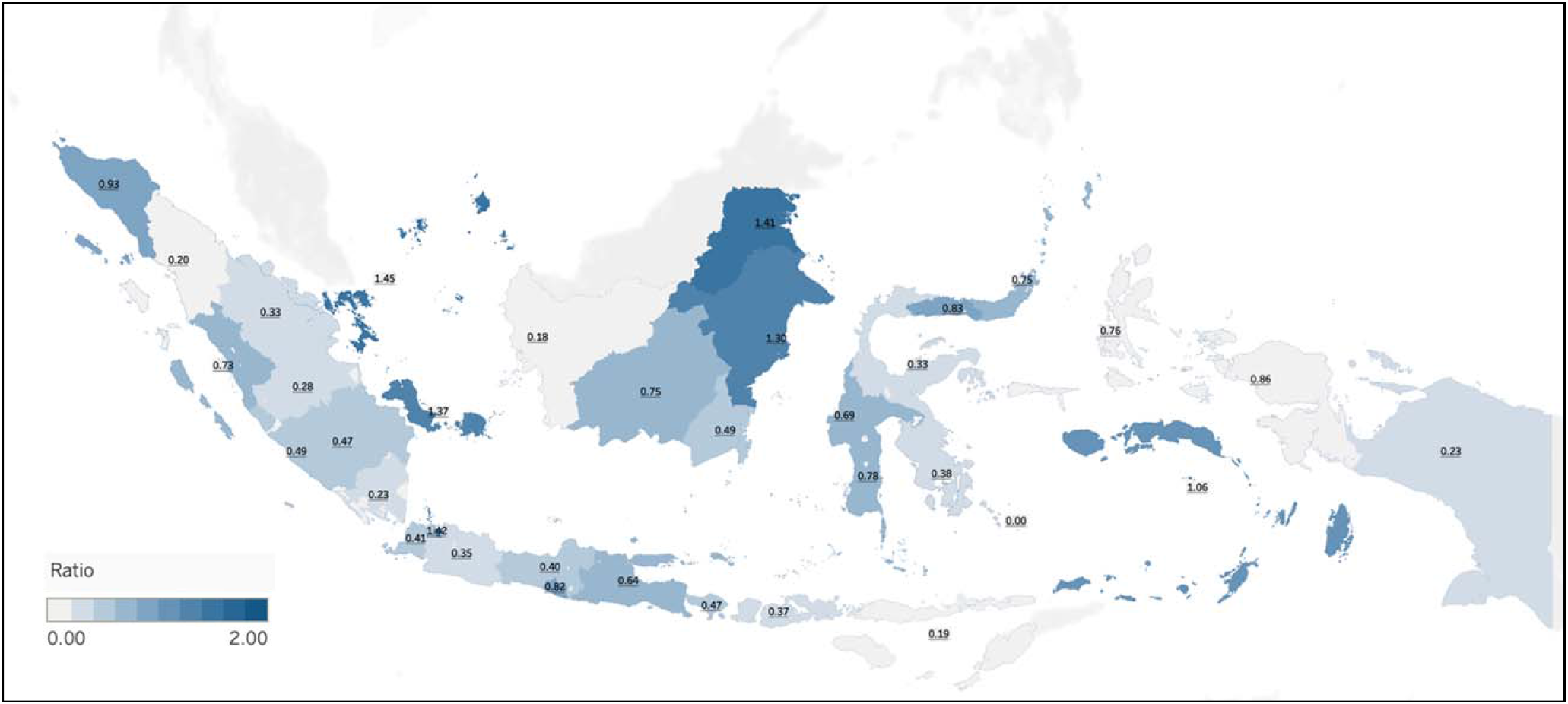
The density of Cathlab per Population in Indonesia

- **Cath lab facility data**

Open-source data for cath labs were unavailable; therefore, we conducted a primary survey from the data of hospital instruments from the Ministry of Health. These data were cross-checked using a primary survey of the regional heart association data and online data mining sourced from hospital facilities’ websites and news of cath lab openings from trusted media. After the primary data were finished, the data was cross-checked with the data from the Ministry of Health, the Indonesian Heart Association, and the Indonesia National Health Insurance. Data that did not match with our sources, or was ambiguous, was further checked by contacting the hospital or the regional heart association. The final cath lab data reflects the situation in Indonesia as of June 2022. Some facilities, especially tertiary and referral hospitals, may have more than one cath lab. Information on the cath labs in every hospital was unavailable publicly. As a compromise, we, therefore, counted the number of hospitals that have a cath lab.

Existing hospital data was also analyzed temporally based on when the PCI facility started operating. Data collection ran from February to June 2022. Facilities that opened after data collection could not be included because of the limited digital data system in Indonesia.

- **Population and Administrative data**

The population data were obtained from the geospatial information system (GIS) data of the Ministry of Internal Affairs. The most recent population estimates were from 30 June 2022. GIS data contain both population and administrative data. Indonesia’s administration level is divided into three levels: province, district, and subdistrict. Province is the term for the first administrative level, and Indonesia has 34 provinces. The second administrative levels are District, that divides into Cities (Kota) and Regencies (Kabupaten). Cities are urban areas with a high population density and a likely higher socioeconomic status, while regencies are suburban to rural regions with a lower socioeconomic status. Indonesia had 416 regencies and 98 cities in 2022.

In addition, we aggregate health worker data into five islands with the densest population: Sumatra, Java, Kalimantan, Bali-Nusa, Sulawesi, and Maluku-Papua. The cath labs are also grouped into public and private hospitals because they serve different target populations. The public hospital focuses on providing services for all levels of society, whether poor or wealthy.

## Density and Gini index

To measure and compare the cath lab maldistribution, we compared the density of the cathlab between areas and used Gini index as inequality measurement. The density was defined as the number of cath labs per population within the same area. The Gini index is a widely recognized measure of economic and healthcare inequality. We were comparing Gini index of cathlab at provincial level Gini Index ranges from 0 to 1, with greater values indicating greater levels of inequality. UNICEF define Gini index as <0·2 corresponds to ideal income equality, 0·2–0·3 corresponds to relative equality, 0·3–0·4 corresponds to a substantial income gap, 0·4–0·5 corresponds to a high-income disparity, and above 0·5 corresponds to severe income inequality. The Gini index was calculated using R in RStudio, “dineq” Package by René Schulenberg.^7^

## Results

### Cath lab growth

The number of cath labs in Indonesia increased significantly from 2017 to 2022. Within six years, 129 new cath lab facilities were added. Increasing the total cath labs from 181 to 310 further contributed to 71·3% of the total cath lab growth. The owner distribution of the cath lab is still dominated by private hospitals, which own 54% of the facilities. In the last six years, 62 private hospitals have new cath labs, whereas public hospitals added 67 [Table 1].

**Table 1.** Indonesia Hospital with cath lab growth from 2017 to 2022.

| Group/Region | Year |  |  |  |  |  | Percentage % |
| --- | --- | --- | --- | --- | --- | --- | --- |
|  | <=2017 | 2018 | 2019 | 2020 | 2021 | Half 2022 |  |
| **Cath lab number (Growth)** | 181 | 221<br>(+40) | 254<br>(+33) | 273<br>(+19) | 294<br>(+21) | 310<br>(+16) | 100% |
| **National insurance-covered** | 87 | 108<br>(+21) | 120<br>(+12) | 129<br>(+9) | 135<br>(+6) | 139<br>(+4) | 44·80% |
| **OWNER GROUPING** |  |  |  |  |  |  |  |
| Public-owned | 80 | 99<br>(+19) | 115<br>(+16) | 125<br>(+10) | 134<br>(+9) | 142<br>(+8) | 45·80% |
| Private-owned | 101 | 122<br>(+21) | 139<br>(+17) | 148<br>(+9) | 160<br>(+12) | 168<br>(+8) | 54·20% |
| **ISLAND GROUP** |  |  |  |  |  |  |  |
| Bali Nusa | 8 | 8 | 8 | 8 | 8 | 9<br>(+1) | 2·90% |
| Java | 127 | 159<br>(+32) | 178<br>(+19) | 188<br>(+10) | 201<br>(+13) | 208<br>(+7) | 67·10% |
| Kalimantan | 6 | 7<br>(+1) | 11<br>(+4) | 14<br>(+3) | 15<br>(+1) | 16<br>(+1) | 5·16% |
| Maluku | 1 | 1 | 1 | 1 | 2<br>(+1) | 2 | 0·65% |
| Papua | 1 | 1 | 1 | 1 | 1 | 1 | 0·32% |
| Sulawesi | 9 | 10<br>(+1) | 13<br>(+3) | 14<br>(+1) | 14 | 18<br>(+4) | 5·81% |
| Sumatra | 30 | 36<br>(+6) | 43<br>(+7) | 47<br>(+4) | 53<br>(+6) | 56<br>(+3) | 18·06% |

In 2018 and 2019, the Growth Rate was up to 33 cath labs per year. When the COVID pandemic occurred, the cath lab growth in Indonesia decreased, dropping to 19 cath labs in 2020 and 21 in 2021. In 2022, we saw an upward trend, with 16 new cath labs opening in the first half of 2022. Java Island still dominates cath lab growth contributing 81 new facilities (62·3% of the total cath labs). Followed by Sumatra with 26 new cath labs (20%), Kalimantan with 10 (7·7%), and Sulawesi with nine (6·9%). There have been no new cath labs in Papua, Bali, and Nusa in the past six years [Table 1].

Table 2 shows that within five years, there were 44 new cath labs in districts previously without one. That is, one-third of the new cath labs were built in districts that did not have one. This increased the districts covered by cath labs from 83 to 113 (21·9% of the total districts). Public-owned hospitals made up the majority of new cath labs in these areas, with 31 of the 44 cath labs. Most of the new cath labs in new districts were built in 2018. After that, the number of cath lab additions in new areas decreased.

**Table 2.** The Distribution of new Hospital with cath labs in districts that did not have one.

| **Hospital Owner** | **Additional Cathlab per Year** |  |  |  |  | **Grand Total** |
| --- | --- | --- | --- | --- | --- | --- |
|  | **2018** | **2019** | **2020** | **2021** | **2022** |  |
| Public | +9 | +7 | +5 | +4 | +6 | 31 |
| Private | +5 | +1 | +3 | +2 | +2 | 13 |
| Total | +14 | +8 | +8 | +6 | +8 | 44 |
| **District-covered** | **83** | **91** | **99** | **105** | **113** |  |

### The density of Hospitals with cath labs and cardiologists at the subnational level

In Java, there are 981 cardiologists and 208 cath labs, the highest levels in Indonesia. Sumatra Island follows this with 216 cardiologists and 56 cath labs. Sulawesi Island has 107 cardiologists and 18 cath labs, while Kalimantan Island, Bali, Lesser Sunda, Maluku, and Papua are almost 50% lower. Kalimantan has 60 and 16, Bali, 52 and 5, and Lesser Sunda 18 and 4 cardiologists and cath labs, respectively. The lowest numbers are in Maluku, which only has six and two, and Papua, with four and one cardiologists and cath labs, respectively [Table 3].

**Table 3.** The distribution of Indonesian cath labs and cardiologists within major islands and provinces.

| Island and Province | Population | Number |  | Density (per million) |  |
| --- | --- | --- | --- | --- | --- |
|  |  | Cardiologist | Cath | Cardiologist | Cath |
| **National** | 269,648,234 | 1,446 | 310 | 5.36 | 1.15 |
| **Lesser Sunda** | **10,713,122** | **18** | **4** | **1.68** | **0.37** |
| West Nusa Tenggara | 5,308,887 | 15 | 3 | 2.8 | 0.57 |
| East Nusa Tenggara | 5,404,235 | 3 | 1 | 0.6 | 0.19 |
| **Bali** | **4,285,815** | **52** | **5** | **12.1** | **1.17** |
| **Java** | **151,629,504** | **981** | **208** | **6.47** | **1.37** |
| DKI Jakarta | 11,207,822 | 327 | 50 | 29.2 | 4.46 |
| Di Yogyakarta | 3,671,818 | 48 | 7 | 13.1 | 1.91 |
| Central Java | 36,947,166 | 111 | 27 | 3 | 0.73 |
| West Java | 47,161,961 | 209 | 53 | 4.4 | 1.12 |
| East Java | 41,002,289 | 229 | 49 | 5.6 | 1.2 |
| Banten | 11,638,448 | 57 | 22 | 4.9 | 1.89 |
| **Kalimantan** | **16,641,765** | **60** | **16** | **3.61** | **0.96** |
| North Kalimantan | 681,766 | 2 | 2 | 2.9 | 2.93 |
| East Kalimantan | 3,770,038 | 24 | 6 | 6.4 | 1.59 |
| West Kalimantan | 5,475,380 | 17 | 2 | 3.1 | 0.37 |
| South Kalimantan | 4,078,949 | 12 | 3 | 2.9 | 0.74 |
| Central Kalimantan | 2,635,632 | 5 | 3 | 1.9 | 1.14 |
| **Maluku** | **3,167,532** | **6** | **2** | **1.89** | **0.63** |
| Maluku | 1,870,414 | 4 | 2 | 2.1 | 1.07 |
| Maluku Utara | 1,297,118 | 2 | 0 | 1.5 | 0 |
| **Papua** | **5,419,011** | **6** | **1** | **1.11** | **0.18** |
| Papua | 4,272,391 | 4 | 1 | 0.9 | 0.23 |
| West Papua | 1,146,620 | 2 | 0 | 1.7 | 0 |
| **Sulawesi** | **19,715,820** | **107** | **18** | **5.43** | **0.91** |
| Gorontalo | 1,193,241 | 5 | 1 | 4.2 | 0.84 |
| Central Sulawesi | 3,012,499 | 3 | 1 | 1 | 0.33 |
| North Sulawesi | 2,657,940 | 26 | 4 | 9.8 | 1.5 |
| South Sulawesi | 8,933,926 | 41 | 10 | 4.6 | 1.12 |
| West Sulawesi | 1,447,712 | 27 | 1 | 18.7 | 0.69 |
| Southeast Sulawesi | 2,470,502 | 5 | 1 | 2 | 0.4 |
| **Sumatra** | **58,075,665** | **216** | **56** | **3.72** | **0.96** |
| Riau Island | 2,012,496 | 7 | 4 | 3.5 | 1.99 |
| Lampung | 9,087,213 | 13 | 6 | 1.4 | 0.66 |
| Riau | 6,015,023 | 16 | 7 | 2.7 | 1.16 |
| West Sumatra | 5,448,684 | 41 | 5 | 7.5 | 0.92 |
| Jambi | 3,534,933 | 12 | 2 | 3.4 | 0.57 |
| Bengkulu | 2,019,339 | 5 | 1 | 2.5 | 0.5 |
| Aceh | 5,301,202 | 21 | 6 | 4 | 1.13 |
| South Sumatra | 8,281,587 | 11 | 7 | 1.3 | 0.85 |
| North Sumatra | 14,938,770 | 82 | 16 | 5.5 | 1.07 |
| Bangka Belitung Island | 1,436,418 | 8 | 2 | 5.6 | 1.39 |

**Table 4.**
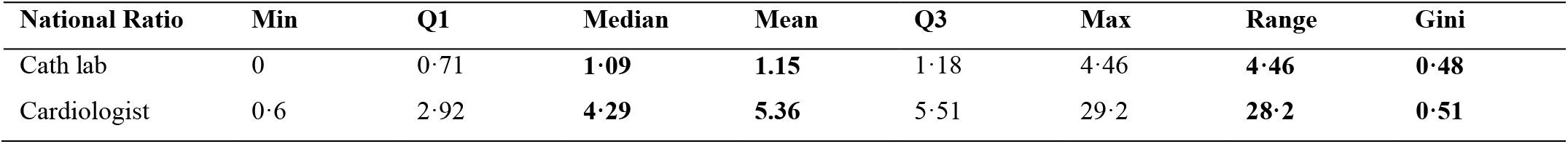
Descriptive statistics of cath lab and cardiologist distribution per million population.

### Cardiologist and Hospital with cath lab distribution

Table 3 shows the number of cardiologists by province. DKI Jakarta has the highest number of cardiologists, with 327, followed by East Java with 229 and West Java with 209. The lowest number is in the provinces of West Papua and North Maluku, with two cardiologists each, followed by Central Sulawesi with three and Maluku with four. The highest density of cardiologists in Indonesia’s islands is found in Bali, with 12·1 cardiologist per million, followed by Java Island with 6·47 and Sulawesi with 5·43. According to the density of cardiologists, DKI Jakarta has the most with 29·2, followed by West Sulawesi with 18·7 and DI Jogjakarta with 13·1. Central Sulawesi has one cardiologist, Papua has 0·9, and NTT has 0·6 for the lowest density.

Regarding cath labs, Java, Bali, and Sumatra have the greatest densities of 1·37, 1·17, and 0·96, respectively. The province with the highest cath lab density is DKI Jakarta, with 4·46 cath labs per million, North Kalimantan came in second with 2·93, and The Riau Island came in third with 1·99. The lowest values were recorded in East Lesser Sunda (0·19 cath lab per million), Papua (0·23), West Papua (0), and North Maluku (0).

## Discussion

### ACS treatment in LMIC

According to the Indonesia Basic Health Research General Statistics, ACS numbers are increasing dramatically in all provinces, with up to 90% increase in prevalence within ten years. The number will continue to rise as the Indonesian population ages. In the next ten years, one-third of the population will be aged over 45, resulting in a higher prevalence of ACS.^8^ In low-and middle-income countries (LMICs), ST-elevation myocardial infarction (STEMI) is one of the most prevalent presentations of ACS. Over 60% of hospitalizations for ACS are due to STEMI in India, and 80%. in China^9–12^

ACS, particularly STEMI, requires prompt intervention and coordination of systems. Creating efficient systems in resource-constrained contexts is challenging. Successful solutions from more developed countries cannot simply be transferred. The challenges include a lack of accessible PCI facilities in urban areas, a shortage of specialists, and inadequate emergency medical services. These structural issues hinder timely and effective care for ACS patients. Indonesia is lagging in this respect, and cath lab utilization is suboptimal. Often, the 24-hour service is unavailable, and providers do not respond quickly to cardiac emergencies. Most LMICs in the Southeast Asian Region, especially Indonesia, only provide 24-hour rescue or primary PCI at the tertiary or national level. ^6,8^

A study in Jakarta, the capital of Indonesia, found that between 2007 and 2013, PCI procedures increased from 24% to 35%, and non-reperfusion patients decreased from 67·1% to 62·8%. Maintaining the expansion of PCI procedures is important as the overall STEMI mortality rate has decreased from 11·7% to 7·5%. Increased cath lab usage indicates progress. However, the problems in other regions of Indonesia are unquestionably worse than in the capital. Especially in regions with no cath labs or where their number and accessibility are limited.^13,14^

### Number and growth of cath labs in Indonesia

In the last five years, the number of cath labs built in Indonesia has increased by up to 40%. This impressive progress is good preparation for the population shift expected in the next ten years. However, the pandemic slowed cath lab growth by up to 50% in 2020 and 2021. This is understandable, but as a result, the Ministry of Health must accelerate cath lab growth over the next decade to meet demand. Java Island has 81 new cath laboratories, two-thirds of the total. Sumatra Island has 26. Papua Island has one cath lab and has not had a new one in five years. According to “The Inverse Care Law,” good medical or social care availability varies inversely with population requirements. The province of West Java, with 53, has the most cath labs in Indonesia, but this is only 1·12 cath labs per million population. EAPCI recommends one cath lab per 500,000–600,000 people or 1·67–2 cath labs per one million. Indonesia’s cath lab ratio grew from 0·68 to 1·14 per million people, but her 272 million population needs at least 450 cath labs to meet the recommendations.

The other concern is 24-hour cath lab services, which remain low in Indonesia. We could not extract the data of facilities that can perform PCI around the clock. EAPCI’s previous study shows other countries’ levels of 24-hour service. The lowest rate was in Egypt (0·2 per million), to the highest in Poland and Belgium (>4 per million). Only a few hospitals in Egypt had catheterization laboratories that offer 24-hour cardiac emergency care, ensuring low rates of primary PCI and structural cardiac interventions in Egypt.^15^ In 2014, Jakarta had only one national referral hospital that provided 24-hour service for primary PCI covered by national health insurance. The amount of data of Hospitals that can handle PCI round the clock is limited. These may be important factors to consider when developing the STEMI network system as a government insurance system must be implemented in hospitals that provide PCI services^18,^.

### Distribution of cath labs in Indonesia

We compared the ratio between regions and the Lorenz curve with the Gini index to measure maldistribution (Figure 4). The density of cath labs in Indonesia ranges from 0·0 in West Papua and Maluku to 4·46 in Jakarta; the median is 1·09. (IQR 0·71–1·18). The wide range of densities shows area disparities. The wide range of ratios is in concordance with Gini Index. Our Analysis shows that the Gini index in 2022 was 0·48. This Indicates a huge national disparity. The Gini index decreased in 2022 compared to 2017, falling from 0·54 to 0·48. Despite a significant increase in cath labs, the small change in the Gini index shows that new cath labs are still concentrated in old cath lab areas.

**Figure 4.**
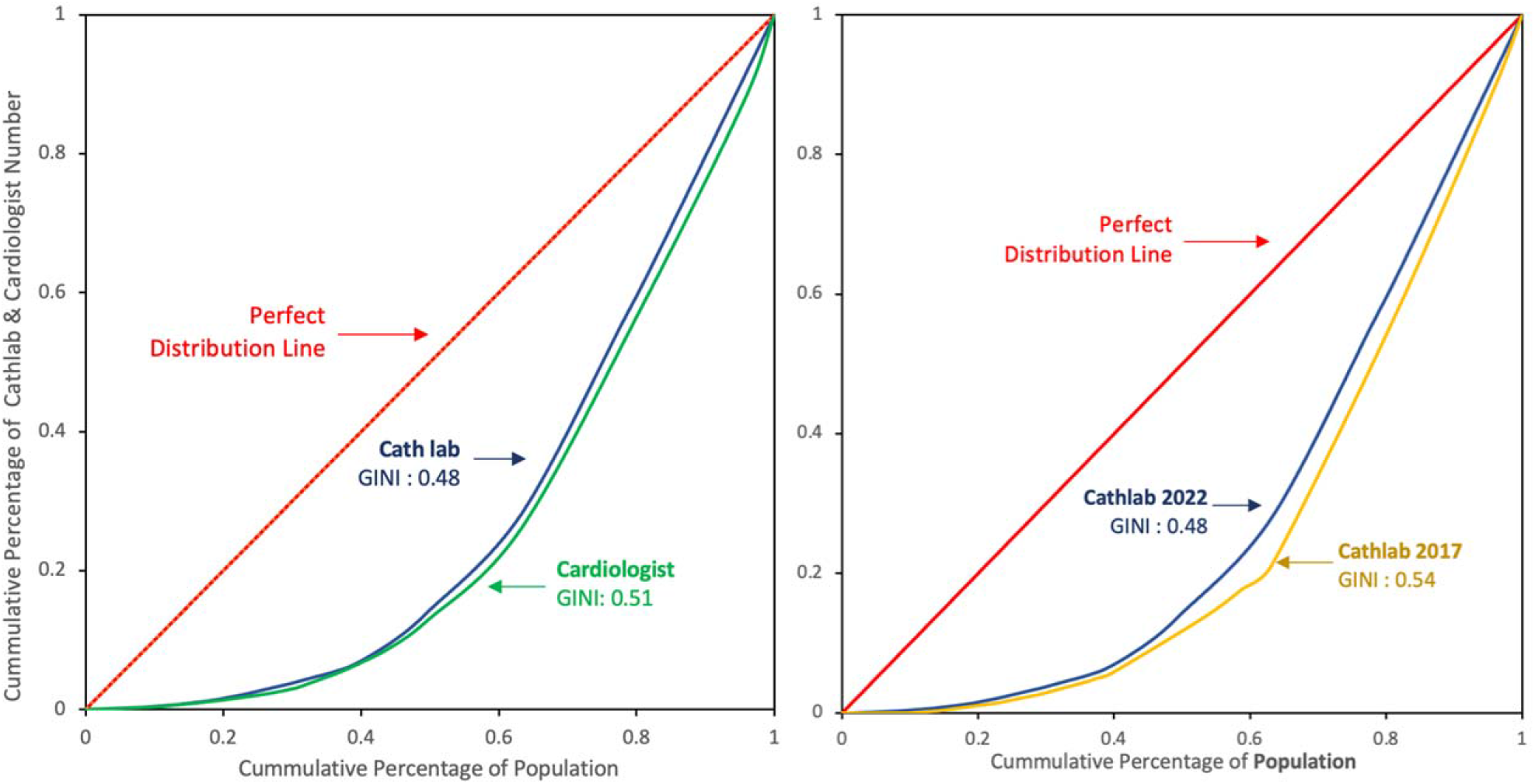
The Lorenz Curve and Gini index of the cath lab and cardiologist distribution,

Public and private hospitals in Indonesia increased cath lab distribution and coverage. Private hospitals own 54% of the cath labs. Public hospitals dominated the provision of first cath labs in the new area. Private hospitals added more cath labs than public hospitals overall, but public hospitals own 31 of the 44 in new areas. Private hospitals often build a cath lab in an area with an existing lab. Building a new cath lab in an area without one is a priority because it will cover more areas. The Ministry of Health advises central and regional public hospitals and maintains a high cath lab ratio.

### Cath lab with insurance coverage

ACS is a catastrophic and expensive disease, typically costing $ 3,000–$ 4,000. As most people in Indonesia are in the middle-to low-income bracket, health insurance is crucial for providing access to cath lab therapy. According to our findings, only 139 cath labs out of 310 (44·8%) have partnered with the national health insurance provider. Even if a hospital has a cath lab, patients are less likely to use one that has not cooperated with insurance.

## RECOMMENDATION

### Policy making for cath lab procurement

Increasing cath lab capacity will improve the outcome for patients. The high Indonesian ACS mortality increases the importance of procuring more cath labs. We can learn from Poland, where cath lab referral services developed five to ten years ago. Poland increased its STEMI patient treatment capacity to more than 16 more advanced European countries^15^. The study also shows that the number of cath labs does not correlate with economic conditions. Greece, with a smaller economy than Denmark, has more cath labs. This shows the importance of the government as a regulator of policy and decision-making in the procurement of cath labs. Increasing resources requires managing procurement effectiveness and efficiency.15,19

### Cath lab mapping

Indonesia must continuously monitor and report using a dashboard system to map the hospitals with cath labs. It can then provide information on how the cath labs are distributed, whether they can do PCIs with their Interventionist and whether they are covered by insurance. Several concerns require systematic investigation. There are information gaps in health services research. Precise mapping of the percentage of the population that can already reach existing thrombolytic/PCI in less than 2 hours would be extremely useful for planning new resources. Such information is mainly unavailable. Geospatial analysis in Russia shows promising results by showing the percentage of the population that can access cath labs promptly and areas that still need a cath lab. Further study of Indonesia cath lab coverage mapping is also required.^18–21^

### Cath lab monitoring

We need to monitor other factors that influence the PCI process at the hospital level. The impact of new programs on patient outcomes and healthcare expenditures should be studied so that the regulator can evaluate cath lab programs.^22^Cath lab regulatory policy needs better data on cath lab capacity in local regions. Regionalized care includes community-wide planning for PCI distribution, emergency programs to diagnose and refer patients to PCI facilities, and regional patient transfer agreements between hospitals.^25,26^

### Further research

Further research may answer questions such as how many people are covered by existing cath labs, how new cath lab procurement influences treatment capacity, service utilization, healthcare outcome and costs, and the optimum placement of new cath labs to cover more population.

## Limitations

This study only examines the existence of the cath lab infrastructure, not whether the cath lab can provide services. Further, we could not investigate patient outcomes because we lacked patient-level data.

## Conclusions

Indonesian ACS treatment is susceptible to its low cathlab density. There is also significant maldistribution, accumulating at several loci. Increasing the number and distribution of cath labs with strategic placement is needed to increase the capacity of emergency cardiovascular services in Indonesia.

## Data Availability

Data are available by requesting an application through email to the corresponding author.

## Acknowledgements

We are grateful to Basuni M.D., and Isman Firdaus M.D. for their time and advice in writing this study.

## Financial Support or Funding

The study was conducted with the researcher’s funds.

## Availability of Data and Materials

Data are available by requesting an application through email to the corresponding author.

